# Invasive pulmonary aspergillosis in critically ill patients with severe COVID-19 pneumonia: results from the prospective AspCOVID-19 study

**DOI:** 10.1101/2020.07.21.20158972

**Authors:** Tobias Lahmer, Silja Kriescher, Alexander Herner, Kathrin Rothe, Christoph D. Spinner, Jochen Schneider, Ulrich Mayer, Michael Neuenhahn, Dieter Hoffmann, Fabian Geisler, Markus Heim, Gerhard Schneider, Roland M Schmid, Wolfgang Huber, Sebastian Rasch

**Affiliations:** Klinik und Poliklinik für Innere Medizin II, Klinikum rechts der Isar der Technischen Universität München, Munich, Germany; Klinik und Poliklinik für Aneasthesiologie und Intensivmedizin, Klinikum rechts der Isar der Technischen Universität München, Munich, Germany; Institut für Mikrobiologie, Immunologie und Hygiene, Technische Universität München, Munich, Germany; Institut für Virologie, Technische Universität München, Munich, Germany

**Author notes:** **Corresponding author**: Lahmer Tobias, MD, Klinik und Poliklinik für Innere Medizin II, Klinikum rechts der Isar der Technischen Universität München, Ismaninger Str. 22, 81675 Munich. equal contribution (Wolfgang Huber, Sebastian Rasch). **Authors’ email addresses**.

**Keywords:** COVID-19, SARS-CoV-2, invasive pulmonary aspergillosis, critically ill, galactomannan, ARDS

## Abstract

**Background:** Superinfections, including invasive pulmonary aspergillosis (IPA), are well-known complications of critically ill patients with severe viral pneumonia. Aim of this study was to evaluate the incidence, risk factors and outcome of IPA in critically ill patients with severe COVID-19 pneumonia.

**Methods:** We prospectively screened 32 critically ill patients with severe COVID-19 pneumonia for a time period of 28 days using a standardized study protocol for oberservation of developement of COVID-19 associated invasive pulmonary aspergillosis (CAPA). We collected laboratory, microbiological, virological and clinical parameters at defined timepoints in combination with galactomannan-antigen-detection from bronchial aspirates. We used logistic regression analyses to assess if COVID-19 was independently associated with IPA and compared it with matched controls.

**Findings:** CAPA was diagnosed at a median of 4 days after ICU admission in 11/32 (34%) of critically ill patients with severe COVID-19 pneumonia as compared to 8% in the control cohort.

In the COVID-19 cohort, mean age, APACHE II score and ICU mortality were higher in patients with CAPA than in patients without CAPA (36% versus 9.5%; p<0.001). ICU stay (21 versus 17 days; p=0.340) and days of mechanical ventilation (20 versus 15 days; p=0.570) were not different between both groups. In regression analysis COVID-19 and APACHE II score were independently associated with IPA.

**Interpretation:** CAPA is highly prevalent and associated with a high mortality rate. COVID-19 is independently associated with invasive pulmonary aspergillosis. A standardized screening and diagnostic approach as presented in our study can help to identify affected patients at an early stage.

**Funding:** None

## Introduction

Since the outbreak of the novel severe acute respiratory syndrome coronavirus 2 (SARS-CoV-2) – associated respiratory disease in December 2019, numerous patients were hospitalized with viral pneumonia and respiratory insufficiency, which was finally designated as the clinical coronavirus disease 2019 (COVID-19) (1). Nearly 5% of the affected COVID-19 patients are critically ill, develop an acute respiratory distress syndrome (ARDS) and need intensive care unit management including mechanical ventilation (1, 2).

Along with other uncertainties during an intensive care unit (ICU) stay, superinfections, including invasive pulmonary aspergillosis (IPA), are well-known complications of severe viral pneumonia in critically ill patients. The association between viral pneumonia and IPA was first reported in a greater cohort during the H1N1 influenza seasons 2009-2011 by Wauters et al. (3). Surprisingly, not only the rate of IPA was higher than suspetced (incidence of 23%) but also nearly half of the IPA patients did not fulfill the classical risk factors of the European organisation for research and tretament of cancer/ mycosis study group (EORTC/MSG) for IPA developement in this cohort (4). These findings could be confirmed by the dutch-belgian mycosis study group and resluted not only in the recognition of influenza as an independent risk factor for IPA devlopement but also in modified definitions and diagnostic criterias for IPA in critically ill patients (5, 6).

Therefore, with the (modified) AspICU algorithm for critically ill patients adapted diagnostic criterias for IPA could be established beside the EORTC/MSG criteria (7). However, the new clinical conditions of COVID-19 patients along with infection control restrictions for biosampling will make the diagnostic procedures and microbiological interpretation for IPA in COVID-19 patients more challenging.

In analogy to what has been reported in critically ill patients with severe influenza associated pneumonia, the aim of our prospective AspCOVID-19 study is to describe the incidence and outcome of COVID-19 associated invasive pulmonary aspergillosis (CAPA) in critically ill patients with severe pneumonia using a standardized screening procedure and assess whether COVID-19 is independently associated with IPA.

## Methods

### Study design and participants

This prospective cohort study was conducted at two tertiary care ICU’s (department of internal medicine and department of anaesthesiology) of the University Hospital of Technical University of Munich, Germany.

Patients (18 years of age or older) with confirmed severe COVID-19 pneumonia (clinical signs, typical laboratory constellation, PCR test for SARS-CoV-2 positive and chest computed tomography (CT) scan with typical signs) who were admitted to the ICU due to acute respiratory failure for more than 48 hours with the need for mechanical ventilation were eligible for study inclusion.

64 COVID-19 negative critically ill patients with ARDS and pneumonia without immunosuppression were included as a retrospective matched control group. Selection criteria for the control cohort were ARDS caused by pneumonia with a corresponding Horowitz Index <150 mmHg as well as comparable sequential organ failure assessment (SOFA) and acute physiology and chronic health evaluation (APACHE II) scores. Exclusion criteria of the control cohort were as follows:

- Patients fulfilling the EORTC/MSG criteria
- Immunosuppression
- mycological evidence from only one specimen from lower repiratory tract and no correlation in broncho-alveolar lavage (BAL) or standard microbiological findings

A total of 347 ICU patients were screened for the retrospective machted control cohort, 283 were excluded (215 didnot fulfill the selection criteria, 67 met the EORTC/MSG criteria, 1 patient was *Aspergillus spp*. colonized). Pregnancy, age younger than 18 years, insufficient available information or lacking written informed consent were general exclusion criterias.

The study was approved by the institutional review board, Klinikum rechts der Isar, TU München (Ref. 149/20S) and registred as a prospective study at ISRCTN (trial registration number 38127). Written informed consent was obtained by the patient or their legal representatives

### Screening for aspergillosis of critically ill patients with COVID-19 and control cohort

Patients of the COVID-19 cohort were prospectively screened in defined time intervals for developement of Covid-19 associated invasive pulmonary aspergillosis (CAPA) following the study protocol (figure 1).

**Figure 1:**
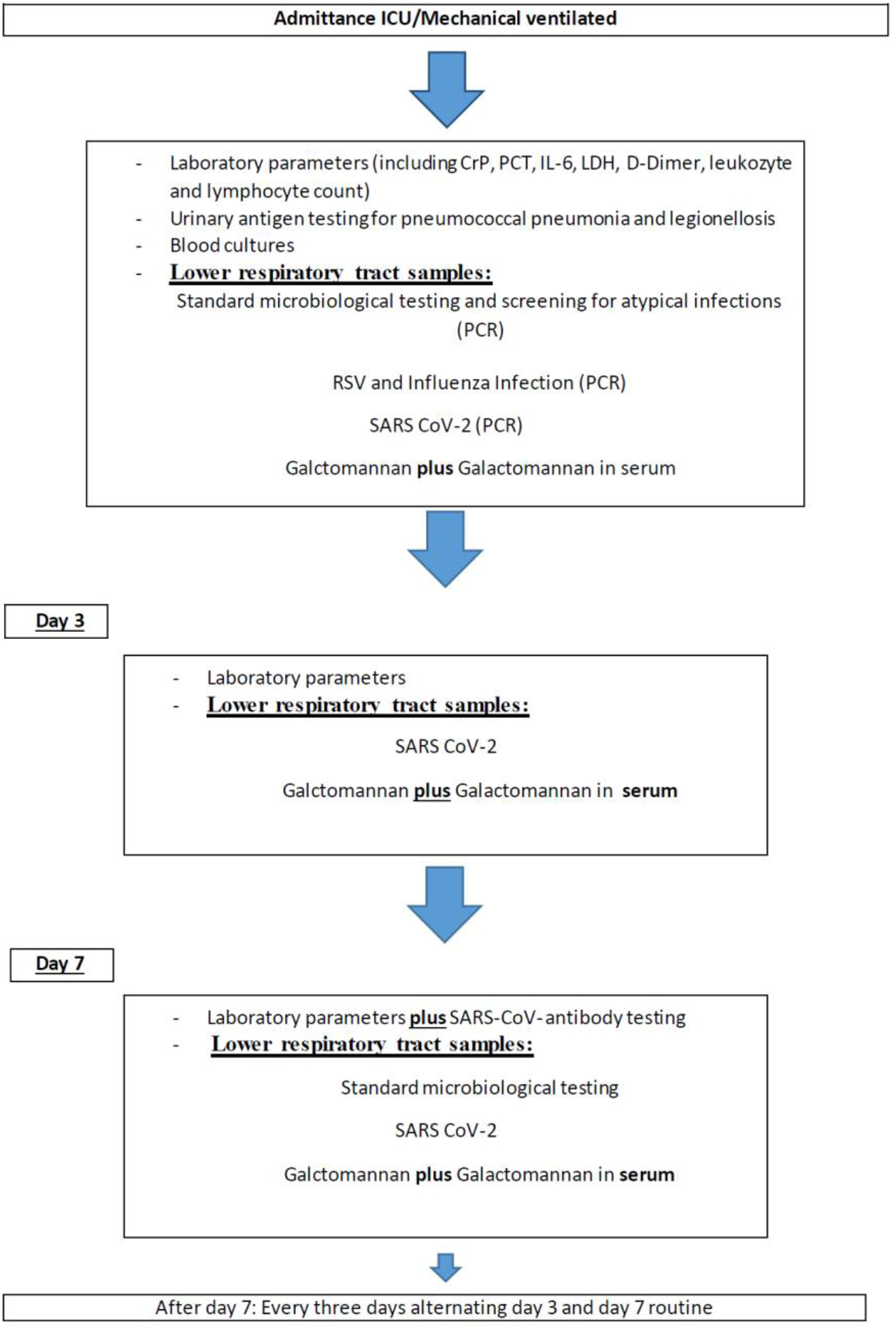
Standardized diagnostic algorithm for patients with suspected or confirmed COVID-19 associated ARDS

In addition standard microbiological, virological and laboratory tests were performed at time intervals summarized in the study protocol (at admission tests were also perforemd before ICU admittance in the emergency department or general ward. If these tests were performed >1 day before ICU admission all test were repeated at ICU admittance). All patients received a chest CTscan before ICU admittance. Testing for atypical pneumonia (using PCR from bronchial aspirates, and pneumococcal antigen from urine) as well as respiratorial syncytial virus (RSV) - and influenza were performed in all patients.

For safety reasons, diagnostic testing of respiratory specimens was performed in accordance to the guidelines of the European society of intensive medicine (ESCIM) with bronchial aspirates (BA) gained by deep bronchial suction with a closed suction system from intubated patients.

Screening for CAPA ended after extubation for all patients. In patients with positive proof of aspergillus galactomannan-antigen (GM) a serum GM follow up examination was performed. Overall observation time for all COVID -19 patients was 28 days.

Results of glactomannan testing from bronchoalveolar lavage (BAL) examinations were used in the control group. These patients were screened for invasive pulmonary aspergillosis once weekly according to our local ICU standard.

### Definitions

The modified AspICU score, developed for the diagnostic assessment of influenza associated IPA, was adapted on COVID-19 patients and used to classify IPA. Putative IPA was assumed in one of the following conditions: cultural growth of *Aspergillus spp*.; GM optical density index (ODI) >0.5 in serum, GM ODI >1 in lower respiratory tract specimen. Clinical signs and radiological signs were in line with the modified AspICU score.

When >1 criterion necessary for CAPA diganosis was not met, these cases were classified as *Aspergillus* colonization.

Every COVID-19 patient fulfilling the mentioned criteria was discussed by a specialist (consultant) for microbiology and a specialist for intensive care medicine (consultant) to ascertain that the criterias for CAPA were appropriate.

## Microbiological and virological methods

Galactomannan (GM) detection (Platelia™ Aspergilllus Ag, Bio-Rad Laboratories, Munich, Germany) was performed in serum samples and in bronchial aspirations (BA) samples gained by deep tracheal suction with a closed suction system from the lower respiratory tract.Primary aerobic microbiological cultures from BA fluid were performed on columbia agar and chocolate agar (prepared culture media, Becton Dickinson, Sparks, MD, United States of America). When growth of *Aspergillus spp*. could be established, it was then sub-cultured on sabouraud-dextrose-agar (Oxoid™ Thermo Fisher Scientific™, Waltham, MA, United States of America) for species identification via macroscopic, microscopic and MALDI-TOF (Bruker Daltronics GmbH, Leipzig, Germany) analysis.

Phenotypic screening for azol-resistance in *Aspergillus spp*. was performed routinely using RPMI (Becton Dickinson, Sparks, MD, United States of America) agar plates supplemented with voriconazol (2mg/L) and itraconazol (4mg/L) and an antifungal-free agar as growth control. Azole susceptible isolates were identified by growth on the antifungal-free agar and absence of growth on plates containing azoles.

## Virological testing

SARS CoV-2 nucleocapsid gene was detected by Taqman RT PCR with the primers 2019-nCoV_N1-F-5’-GAC CCC AAA ATC AGC GAA AT-3’, 2019-nCoV_N1-R 5’-TCT GGT TAC TGC CAG TTG AAT CTG-3 and the probe 2019-nCoV_N1-P 5’-FAM-ACC CCGTAC GTT TGG TGG ACC-BHQ1-3’ (https://www.cdc.gov/). Positive results wereconfirmed with another pair of N gene primers and-probe. IgG and IgM antibodies directed at SARS-CoV were detected with the iFlash 1800Chemiluminescence Immunoassay Analyzer (YHLO, China).

### Statistical analysis

Statistical analysis was performed using IBM SPSS Statistics 25 (SPSS Inc, Chicago, Illinois, USA). Samples were checked for normal distribution using the Shapiro-Wilk test. Descriptive data of normally distributed parameters are presented as mean ± standard deviation and as median and range for non-parametric parameters. The Mann-Whitney-U and Kruskas-Wallis tests were used to analyze non-parametric variables and the t-test and a one-way analysis of variances (ANOVA) to analyze variables with normal distribution. To compare qualitative parameters, chi-square test and in small samples (expected frequency of test variable less than 5) Fisher’s exact test was used. Probabilities are displayed as odds ratio (OR) with 95% confidence interval (CI). All statistical tests were two-sided with a level of significance (p-value) of 5%. Binary logistic regression models were used to identify risk factors for aspergillosis. Factors with a p-value below 0.05 in univariate analysis were included in the regression models. To control the false discovery rate after multiple testing we adjusted the level of significance to p=0.015 by the Benjamini-Hochberg procedure for qualitative parameters in the COVID-19 positive patient cohort.

Based on previous studies, sample size was estimated as follows : Assuming 5% of invasive pulmonary aspergillosis in an overall patient population and 20% (estimated from influenza patients) in critically ill COVID-19 patients with severe pneumonia, we needed 32 patients in the COVID-19 cohort and 64 patients in the control cohort to gain a power of 0,786.

## Results

Between March and April 2020, 32 patients with severe COVID-19 associtaed pneumonia were prospectively included in the AspCOVID-19 study. Influenza and respiriatory syncytial virus were negative in all included patients.

Basic patient characteristics of the COVID-19 cohort are summarized in table 1. All patients in the COVID-19 cohort were SARS -CoV-2 PCR positive at ICU admission.

**Table 1:** Baseline characteristics of the COVID-19 cohort

|  | **COVID Cohort**<br>**(n=32)** | **With CAPA**<br>**(n=11)** | **Without CAPA**<br>**(n=21)** | **p value** |
| --- | --- | --- | --- | --- |
| **Baseline characteristics** |  |  |  |  |
| Median age , years (range) | 69.5 (27-84) | 72 (58-84) | 59 (27-84) | 0.065 |
| Male sex (%) | 23 (72) | 7 (63) | 16 (76) | 0.195 |
| Mean APACHE II (SD) | 18±4 | 22±3 | 17 ±3 | **<0.001** |
| Mean SOFA (SD) | 10±3 | 12±2 | 9 ±3 | **0.003** |
| **Risk factors n (%)** |  |  |  |  |
| COPD | 3 (10) | 2 (18) | 1 (5) | 0.266 |
| Asthma | 1 (3) | 0 (0) | 1 (5) | 1.00 |
| DM type 2 | 8 (25) | 3 (27) | 5 (24) | 1.00 |
| Art. hypertension | 21 (65) | 7 (63) | 14 (66) | 1.00 |
| Coronary heart disease | 2 (6) | 1 (9) | 1 (5) | 1.00 |
| CKD | 5 (16) | 1 (9) | 4 (19) | 0.637 |
| Atrial fibrillation | 5 (16) | 1 (9) | 4 (19) | 0.637 |
| **COVID-19 specific characteristics** |  |  |  |  |
| SARS-CoV-2 PCR positive at admission | 32 | 11 | 21 | 0.670 |
| Symptoms before hospital admission, days (SD) | 4± 2 | 3±1 | 4±2 | 0.080 |
| Mean days at general ward, (SD) | 2±2 | 1±2 | 7±2 | 0.510 |
| Number of direct ICU admissions | 20 | 7 | 13 | 0.235 |
| Influenza/RSV PCR negative | 32 | 11 | 21 | - |
| Median days of fever, (range) | 8 (3-12) | 9 (3-15) | 8 (4-13) | 0.260 |
| **ICU data** |  |  |  |  |
| Mechanical ventilation | 32 | 11 | 21 | 0.670 |
| Mechanical ventilation days (range) | 16 (3-28) | 20 (8-28) | 15 (3-28) | 0.570 |
| Need for vasopressors (%) | 32 (100) | 11 (34) | 21 (66) | 0.490 |
| Renal replacement (%) | 9/32 (28) | 6/11 (55) | 3/21 (14) | 0.380 |
| Broad-spectrum antibiotics | 32 | 11 | 21 | 0.670 |
| **Outcome data** |  |  |  |  |
| Median days of ICU stay (range) | 18 (5-28) | 21 (9-28) | 17 (5-28) | 0.222 |
| ICU mortality (%) | 6 (19) | 4 (36) | 2 (9,5) | **<0.001** |
| <u>28 days status :</u> |  |  |  |  |
| - ICU | 11 | 4 | 7 | 0.145 |
| - Hospital | 5 | 2 | 4 | 0.266 |
| - Discharge | 10 | 1 | 8 | **0.025** |

All patients received a chest CT scan before ICU admittance with typical signs for COVID-19 pneumonia but no specific signs for IPA. Results of portable chest x-ray controlls during ICU-stay revealed only unspecific infiltrates. Broad-spectrum antibiotics were initiated in all critically ill patients. Standardized laboratory parameters according to the study protocol included C reactive protein, procalcitonin, interleukin 6, lactat dehydrogenases, d-Dimer, leukocyte coount, and lymphocyte count. No statistical significance could be observed between COVID-19 patients with and without CAPA except for Interleukin-6 (median 259, range 28-793 versus median 118, range 12-234; p=0.013). Laboratory parameters for CAPA survivors and non-survivors are listed in table 2.

**Table 2:** Characteristics of CAPA survivors and non-survivors

|  | CAPA Survivors<br>(N=7) | CAPA Non-Survivors<br>(N=4) | p value |
| --- | --- | --- | --- |
| **Baseline and ICU characteristics** |  |  |  |
| Mean age , years (SD) | 74±8 | 70±8 | 0.889 |
| Male sex (n, %) | 5 (71) | 2 (50) | 0.413 |
| Mean APACHE II (SD) | 21±2 | 24±2 | 0.134 |
| Mean SOFA (SD) | 11±1 | 14±1 | **0.026** |
| Median days of ICU stay (IQR) | 23 (14-27) | 17 (12-22) | 0.276 |
| Mechanical Ventilation days (IQR) | 21 (11-25) | 17 (12-22) | 0.089 |
| **Diagnostics** |  |  |  |
| Positive BA galactomannan | 7 | 4 | - |
| Positive serum galactomannan | 0 | 4 | - |
| BA culture positive (n) | 5 | 4 | - |
| - *Asp. fumigatus* | 5 | 4 | - |
| AspICU criteria |  |  |  |
| - proven | 0 | 0 |  |
| - putative | 7 | 4 |  |
| - Coloniser | 3 | 0 |  |
| Mean Days from Intubation till CAPA diagnostic (IQR) | 4 (1-7) | 4 (1-7) |  |
| Mean antimycotic treatment days (SD) | 21±3 | 17±4 |  |
| Initial treatment (n) |  |  |  |
| - Voriconazole | 4 | 1 |  |
| - Isavuconazole | 0 | 1 |  |
| - Liposomal amphotericin B | 3 | 2 |  |
| <u>Median BA GM (IQR):</u> |  |  |  |
| - Day 1 | 4.6 (4.6-6.7) | 6.3 (6.3 -7.4) | 0.182 |
| - Day 3 | 2.5 (1.8-6.4) | 3.5 (1.8-6.1) | 0.958 |
| - Day 7 | 5.1 (3.8-7.4) | 4.5 (2.8-6.2) | 0.682 |
| - Day 10 | 3.1 (1.6-4.4) | 4.3 (1.2-5.8) | 0.928 |
| - Day 14 | 2.8 (1.2-4.8) | 5.7 (5.6-5.8) | 0.118 |
| - Day 17 | 2.4 (2.1-3.5) | 2.9 (2.9 -3.2) | 0.976 |
| - Day 20 | 2.5 (1.5-3.5) | 5.5 ( 5.5 -6.7) | 0.097 |
| - Day 23 | 2.5 (1.5-3.5) | - | - |
| - Day 27 | <0.5 | - | - |
| <u>Mean Serum GM</u> |  |  |  |
| - Day 1 | <0.5 | 1.5 | - |
| - Day 3 | <0.5 | 1.2 | - |
| - Day 7 | <0.5 | 1.5 | - |
| - Day 10 | <0.5 | 0.8 | - |
| - Day 14 | <0.5 | 1.3 | - |
| - Day 17 | <0.5 | 0.8 | - |
| - Day 20 | <0.5 | - | - |
| - Day 23 | <0.5 | - | - |
| - Day 27 | <0.5 | - | - |
| SARS-CoV-2 PCR positive at diagnosis of CAPA (n) | 7 | 4 |  |
| Mean SARS-CoV-2 PCR positive days (IQR) | 12 (9-20) | 17 | **<0.001** |
| Number of patients with SARS-CoV-2 antibodies | 7 | 0 | **<0.001** |
| Initial Horowitz index and follow up after initiation of antimycotics: |  |  |  |
| -Day 1 |  |  |  |
| -Day 3 | 118 | 77 | **0.036** |
| -Day 7 | 145 | 91 | **0.030** |
|  | 190 | 104 | **0.048** |
| <u>Interleukin 6(SD):</u> |  |  |  |
| - Day 1 | 344±248 | 228±195 | 0.659 |
| - Day 3 | 346±323 | 1453±688 | **0.006** |
| - Day 7 | 174±136 | 1970±824 | **0.002** |
| - Day 10 | 142±65 | 1119±815 | **<0.001** |
| - Day 14 | 164±56 | 1001±567 | **0.038** |
| - Day 17 | 83±57 | 189±24 | 0.080 |
| - Day 20 | 70±8 | 345±0 | **0.034** |
| - Day 23 | 80±18 | - | - |
| - Day 27 | 65±43 | - | - |
| <u>LDH (SD):</u> |  |  |  |
| - Day 1 | 529±261 | 637±462 | 0.243 |
| - Day 3 | 388±120 | 480±90 | 0.777 |
| - Day 7 | 331±99 | 1104±278 | **0.004** |
| - Day 10 | 294±54 | 376±160 | **0.012** |
| - Day 14 | 341±76 | 415±129 | 0.260 |
| - Day 17 | 311±60 | 281±0 | 0.711 |
| - Day 20 | 274±16 | 1540±0 | 0.645 |
| - Day 23 | 242±50 | - | - |
| - Day 27 | 213±30 | - | - |

COVID-19 associated invasive pulmonary aspergillosis (CAPA); bronchial aspirates (BA); acute physiology and chronic health evaluation (APACHE II); sequential organ failure assessment (SOFA); galactomannan (GM); lactat dehydrogenases (LDH)

In total, 11 (34%) of 32 critically ill COVID-19 patients fulfilled the modified invasive pulmonary aspergillosis definition for putative invasive pulmonray aspergillosis (see above). Three patients did not meet the modified criteria; these were defined as colonised and excluded from the study. Median time till diagnosis of invasive pulmonary aspergillosis was 4 days (range 1-7) after ICU admission and intubation.

In the COVID-19 Cohort, mean age (mean: 72 versus 60 years; p=0.003), APACHE II score (mean: 23 versus 17; p= 0.027) and ICU mortality were higher in patients with CAPA than in patients without CAPA (36% versus 9.5%; p<0.001).

ICU stay (21 days (range 9-28) versus 17 days (range 5-28); p=0.340) and mechanical ventilation days (20 days (range 8-28) versus 15 days (range 3-28); p=0.570) were not different between COVID-19 patients with and without CAPA.

Dividing the COVID-19 cohort into patients of CAPA survivors (n=7) and CAPA non-survivors (n=4) the following differences could be observed:

SOFA score (mean: 11 versus 14; p= 0.026), intially Horowitz Index and follow up Horrowitz Index after antimycotic initiation were significantly different for CAPA non survivors as compared to CAPA survivors (day 1: 77 versus 118; p=0.036; day 3: 91 versus 145; p=0.030; day 7: 104 versus 190; p=0.048). Mechanical ventilation days (21 days (range 11-25) versus 17 days (range 12-22); p=0.089) were not significantly different.

In 9 (82%) of the 11 patients with IPA, cultural growth of *Aspergillus spp*. Could be established revealing *Asp. fumigatus* in all cases. GM-ODI in all bronchial aspirate (BA) specimes of CAPA patients was >1 in repeated measurements. Positive serum GM could only be observed in the CAPA non-survivors group. All patients received antimycotic therapy. Therapy monitoring using BA GM revealed decreasing GM levels during observation time in the CAPA survivors but not in the CAPA non survivors (see table 2).

SARS-CoV-2 PCR was positive in all CAPA patients at ICU admittance. Mean days of SARS CoV-2 positivity were significantly shorter in CAPA survivors than in CAPA non survivors (12 (range 9-20) versus 17 days; p<0.001). None of the CAPA non survivor patients developed antibodies against SARS CoV-2 in contrast to all CAPA surviors (p<0.001).

After 28 days significantly more patients without CAPA were discharged from the hospital (8 versus 1; p=0.025), no differences could be observed between ICU and general ward stay at day 28 (see table 1).

In the COVID-19 cohort 11 (34%) were diagnosed with IPA in contrast to 5 (8%) out of 64 patients in the COVID negative control cohort (see table 3).

**Table 3:** Baseline characteristics of the COVID-19 negative control cohort

|  | COVID negative Cohort (n=64) | Without IPA (n=59) | With IPA (n=5) | p value |
| --- | --- | --- | --- | --- |
| **Baseline characteristics** |  |  |  |  |
| Mean age , years (SD) | 68±15 | 68±16 | 58±12 | 0.154 |
| Male sex (%) | 44 (65) | 40 (65) | 4 (80) | 0.160 |
| Mean APACHE II (SD) | 20±3 | 19±2 | 25±4 | **<0.001** |
| Mean SOFA (SD) | 10±2 | 10±2 | 13±2 | **0.002** |
| **Risk factors** |  |  |  |  |
| **n (%)** |  |  |  |  |
| COPD | 13 (20) | 11 (18) | 2(40) | 0.119 |
| Asthma | 5 (7,5) | 5(8) | 3(60) | 0.145 |
| DM type 2 | 22 (33) | 22 (36) | 5 (100) | **<0.001** |
| Art. hypertension | 25 (37) | 24 (39) | 1 (20) | **0.003** |
| Coronary heart disease | 32 (50) | 31 (50) | 1 (20) | **<0.001** |
| CKD | 19(38) | 16 (26) | 3 (60) | 0.368 |
| Atrial fibrillation | 3 (5) | 3(5) | 4 (80) | 0.286 |
| Immunosuppression | 0 | 0 | 0 |  |
| Liver cirrhosis | 13(20) | 11 (18) | 2 (40) | 0.119 |
| Pancreatitis | 9 (14) | 8 (13) | 1 (20) | 0.470 |
| **ICU data** |  |  |  |  |
| Mechanical ventilation (n) | 64 | 59 | 5 | 0.156 |
| Mechanical Ventilation days (IQR) | 17 (7-38) | 16 (6-38) | 21(7-29) | 0.266 |
| Need for vasopressors (n,%) | 64 (100%) | 59 (100%) | 5 | 0.123 |
| Renal replacement (n,%) | 25/64 (39) | 20/59 (34) | 5/5 (100) | **<0.001** |
| **Outcome data** |  |  |  |  |
| Median days of ICU stay (IQR) | 18 (7-38) | 18 (7-38) | 23 (7-35) | 0.190 |
| ICU mortality (n, %) | 37 (55) | 34 (55) | 3 (60) | 0.820 |
| **Diagnostics** |  |  |  |  |
| Median BAL Galactomannan (range) |  | <0.5 | 2.9 (1.8-6.4) |  |
| Median serum galctomannan (range) |  | <0.5 | 0.9 (0.7-1.5) |  |
| BAL culture positive (n) |  |  | 3 |  |
| AspICU criteria (n) |  |  |  |  |
| - Proven |  |  | 0 |  |
| - Putative |  |  | 5 |  |
| - Coloniser |  |  | 0 |  |
| *Asp. fumigatus* |  |  | 3 |  |

Abbreviations:

### Abbreviations

Invasive pulmonary aspergillosis (IPA); acute physiology and chronic health evaluation (APACHE II);sequential organ failure assessment (SOFA); chronic obstructive pulmonary disease (COPD); diabetes mellitus type 2; chronic kidney disease; intensive care unit (ICU), broncho alveolar lavage (BAL)

The basic characteristics of both groups are presented in table 4.

**Table 4:** Patient characteristics of COVID-19 and the control cohort

|  | **COVID-19 cohort**<br>**(n=32)** | **Control cohort**<br>**(n=64)** | **p value** |
| --- | --- | --- | --- |
| **Baseline characteristics** |  |  |  |
| Mean age , years (SD) | 65±15 | 68±15 | 0.372 |
| Male sex (%) | 23 (72) | 44 (65) | 0.753 |
| Mean APACHE II (SD) | 18±4 | 20±3 | 0.103 |
| Mean SOFA (SD) | 10±3 | 10±2 | 0.466 |
| **Comorbidities** |  |  |  |
| **n (%)** |  |  |  |
| COPD | 3 (10) | 13 (20) | 0.175 |
| Asthma | 1 (3) | 5 (7,5) | 0.660 |
| DM type 2 | 8 (25) | 22 (33) | 0.350 |
| Art. hypertension | 21 (65) | 25 (37) | **0.014** |
| Coronary heart disease | 2 (6) | 32 (50) | **<0.001** |
| CKD | 5 (16) | 19(38) | 0.134 |
| **ICU data** |  |  |  |
| Mechanical Ventilation days (range) | 16 (3-28) | 17 (7-38) | 0.889 |
| Median days of ICU stay (IQR) | 18 (5-28) | 18 (7-38) | 0.929 |
| ICU mortality (n, %) | 6 (19) | 37 (55) | **<0.001** |

acute physiology and chronic health evaluation (APACHE II); sequential organ failure assessment (SOFA); chronic obstructive pulmonary disease (COPD); diabetes mellitus type 2; chronic kidney disease; intensive care unit (ICU)

In the total patient cohort including the negative controls COVID-19 (11/16 versus 21/80, p=0.001), higher APACHE II score (18.7±3.1 versus 23.0±3.1; p<0.001), higher SOFA score (median 10, range 4-17 versus median 12, range 10-15; p<0.001) and coronary heart disease (2/16 versus 32/80, p=0.036) were associated with IPA.

To assess whether COVID-19 was independently associated with IPA, a binary logistic regression analysis was performed. This analysis confirmed an independent association between COVID-19 and IPA. Also a higher APACHE II score was independently associated with IAP (Figure 2A).

**Figure 2:**
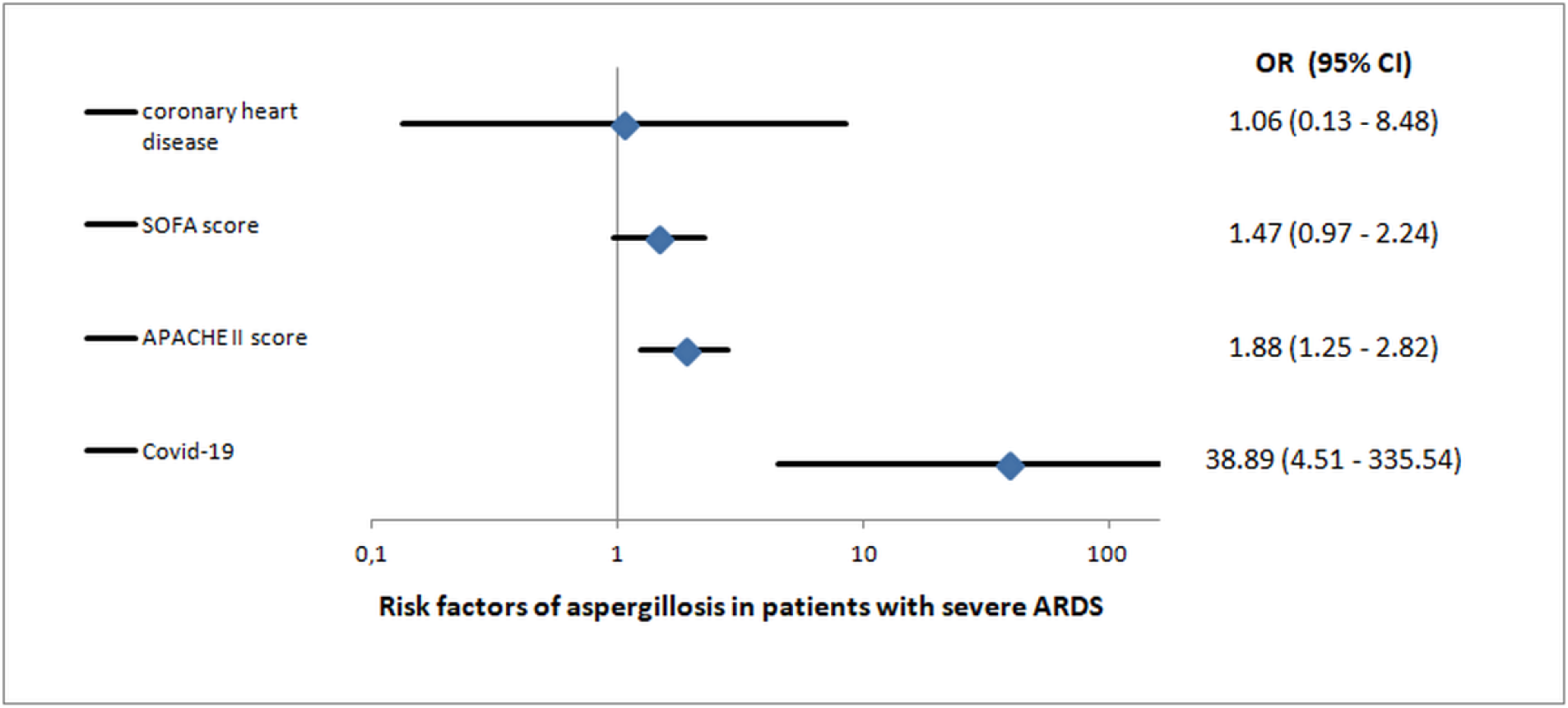
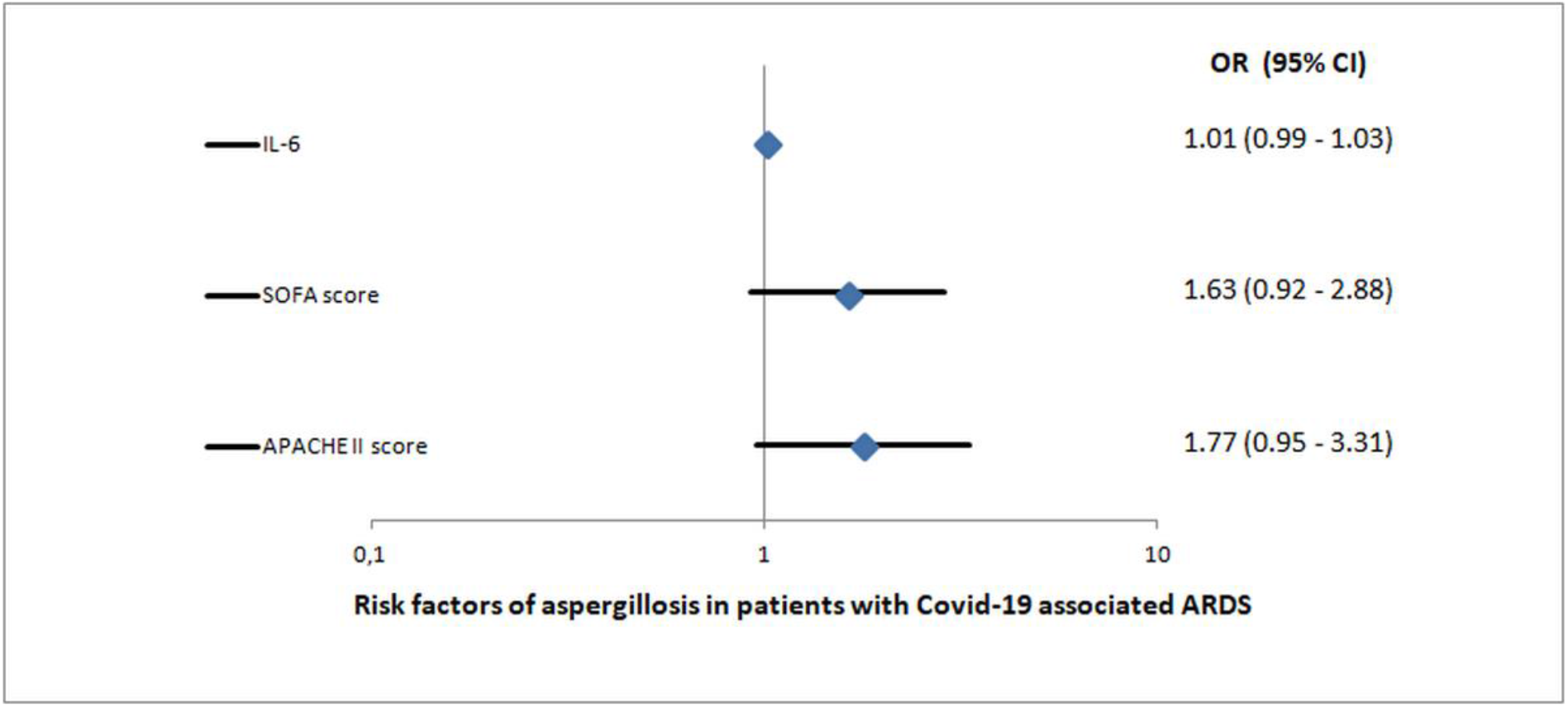
Results of regression models displayed by Forest Blots: A) Risk factors of invasive pulmonary aspergillosis in total patient cohort (COVID-19 patients and controls) B) Risk factors of invasive pulmonary aspergillosis in patients with COVID-19 (OR: odds ratio; 95% CI: 95% confidence interval; IL-6: interleukin −6)

Regarding the COVID-19 cohort, none of the univariate significant parameters were independently associated with IAP in the logistic regression analysis (Figure 2B).

Details of the regression models are presented in supplementary table 1.

**Supplementary table 1:** Details of regression models with IAP as dependent variable in the total patient cohort and the COVID-19 positive patient cohort.

| Total patient cohort including COVID-19 positive and negative patients (n=96) |  |  |  |  |
| --- | --- | --- | --- | --- |
| Independent variable | p in univariate analysis | Exp(B) | 95% CI | p in regression analysis |
| COVID-19 | 0.001 | 38.9 | 4.5 – 335.5 | 0.001 |
| APACHE II score | <0.001 | 1.9 | 1.3 – 2.8 | 0.002 |
| SOFA score | <0.001 | 1.5 | 1.0 – 2.8 | 0.07 |
| Coronary heart disease | 0.036 | 1.1 | 0.1 – 8.5 | 1.0 |
| COVID-19 positive patient cohort (n=32) |  |  |  |  |
| Independent variable | p in univariate analysis | Exp(B) | 95% CI | p in regression analysis |
| APACHE II score | <0.001 | 1.8 | 1.0 – 3.3 | 0.072 |
| SOFA score | 0.003 | 1.6 | 0.9 – 2.9 | 1.0 |
| IL-6 at ICU admission | 0.012 | 1.0 | 0.99 – 1.03 | 0.248 |

## Discussion

As far as we know, up to know our study is the largest prospective study on incidence, risk factors and outcome of IPA in critically ill patients with severe COVID-19 pneumonia.Moreover, our study shows, that COVID-19 is an independent risk factor for IPA.

In comparison to our control cohort, COVID-19 increased the risk of developing an IPA from 8% to 34% and CAPA is associated with a mortality rate of 36%. These findings are as high as previously shown mortality rates, espcially reported from patients with severe influenza pneumonia.

Although, typical risk factors for COVID-19 e.g. arterial hypertension, diabetes mellitus type 2 or coronary heart disease were also significantly more frequent in our COVID-19 cohort, these risk factors were not associated with an increased risk of developing CAPA (8).

Moreover, none of these patients fulfill the typical EORTC/MSG host factor criteria, and only a few COVID-19 patients (4/32) had a history of a chronic pulmonary disease prior to the infection with SARS-CoV-2.

In fact, most patients with COVID-19 presented with mild flu-like symptoms, but up to 15% of the affected patients required assisted oxygenation and 5% of them deteriorated towards a severe ARDS as presented in our study cohort (8). Data on intrinsic risk factors which may predispose to severe ARDS-in COVID-19 patients are sparse. Only small pathological studies of patients with severe COVID-19 associated ARDS, report a typical diffuse alveolar damage combined with intra-alveolar neutrophilic infiltration and vascular congestion, which is interpreted as an acute phase component (9, 10). Therefore, SARS-CoV-2 might trigger an imbalanced immune response resulting in a ‘cytokine stom’ and extensive pulmonary inflammation (11). If the proposed mechanism causing lung injury is a consequence of the described pathological findings or vice versa has to be investigated in further studies. Taken together, it could be assumed that these findings, similary reported in SARS, may lead to an impaired mucociliary activity stimulated by immune cell dysfunction and immune system dysregulation which paves the way for secondary infections (9,11).

It has been shown in several studies including ours, that these super-infections have a negative impact on the outcome of affected patients. (12, 13). The overall incidence of IPA is in line with reported rates for critically ill patients varying between 1 and 7% (12, 13). Furthernore our data increases the awareness of IAP as complication in patients with COVID-19 associated ARDS and helps clinicians to establish standardized screening methods for invasive pulmonary aspergillosis and to early identify high risk patients.

In this study we used the modified AspICU score for diagnosis of CAPA in combination with standardized time intervals of screening. As clinical criteria such as ongoing fever, dyspnea or worsening respiratory insufficiency are also typical of COVID-19 and radiological findings in non-neutropenic patients in most cases do not allow to discrimante typical mycological findings from COVID-19, the diagnosis of CAPA is mostly based on mycological criterias (14).

GM-detection in BAL is a valid test to confirm or rule out IPA with a sensitivity and specificity of approximatley 90% using an ODI-cut-off of ≥0.8 (15).

Due to safety concerns regarding - aerosolization and surface stability of SARS-CoV-2, only bronchial aspirates from deep bronchial suction via a closed system were used for diagnostics as recommend by ESCIM guidelines (16, 17). If the mentioned ODI cut-off for BAL is also reliable for BA specimenis is still a matter of debate. Moreover, increasing the ODI cut-off does not necessarily increase the sensitivity and specificity as reported in some studies (18). Thus, as recommended and used in several studies we used a galactomannan ODI cut-off of >1 for BA equivalent to BAL specimens. Following these considerations we found putative IPA in 34% of our critically ill COVID-19 patients, which is nearly a similar rate to what has been observed in patients with severe influenza pneumonia. Additionally, the median BA GM ODI of all patients within the CAPA cohort was 5.4 (range 1.8-7.4). Moreover, the cultural growth of *Aspergillus spp*. From BA-specimens in 80% of the CAPA patients together with serum GM-detection with ODI >0.5 in 4 CAPA patients strongly emphazise that CAPA is a relevant complication in severe COVID-19 pneumonia. Mean timepoint of CAPA diagnosis was day 4 of ICU stay, which seems early. This may be explained by the fact that all patients already have had reported severe COVID-19 symptoms for several days prior to hospital admission. Antimycotic therapy was initiated in all patients with putative IPA according to recent guidelines (19). The decreasing BA galactomannan levels and increasing Horovitz indexes during therapy suggest an effect of the specific antmycotics beside general ARDS management in the CAPA survivors group. Regarding the high observed incidence of CAPA, the role of antifungal prophylaxis should be further studied.

Nevertheless, there was no difference in median ventilation and ICU days between the COVID-19 cohort with and without CAPA, which may be explained by the severity ofCOVID-19 itself. CAPA patients were significantly older, had a higher APACHE II score and a higher mortality rate as mentioned above.

However, some findings in the CAPA non survivors group were striking. Increased serum interleukin-6 and LDH levels could be associated with worse outcome in COVID-19 patients (20). These findings could be confirmed in the CAPA non-survivors with significantly elevated serum levels, especially of interleukin 6. Moreover, the role of SARS-CoV-2 not only in the pathophysiology of lung injury but also in enhancing an ongoing infection have to be investigated in further studies. In our cohort all CAPA survivors cleared the detectable SARS-CoV-2 RNA and generated IgG antibodies during their ICU stay in contrast to CAPA non survivors with virus persistence until death.

Our study had some limitations. First, it is a single center experience and, confounding cannot be ruled out. However, this is the first prospective study evaluating a standardized screening tool in patients with severe COVID-19 pneumonie in comparison to a retrospective control cohort.

Secondly, the usage of BA instead of BAL is novel and has not been evaluated in larger studies. Given the observations in our study cohort, showing that diagnosis was confirmed in several follow up examinations and by cultures, as well as the special circumstances in COVID-19 patients, we believe, that BA in mechanical ventilated patients gained through deep bronchial suction, are a suitable alternative for GM-testing.

Finaly, due to the novelty of COVID-19 there is still limited information about pathophysiology and clinical characteristics particularly in critically ill patients. Therfore, further studies are needed to analyse out risk profiles for development of CAPA.

In conclucion, in critically ill COVID-19 patients, Covid-19 associated invasive pulmonary aspergillosis is highly prevalent and associated with a high mortality rate. COVID-19 and a high APACHE II score are independently associated with invasive pulmonary aspergillosis. A standardized screening and diagnostic approach as presented in our study can help to identify out affected patients at an early stage.

## Data Availability

The authors confirm that, for approved reasons, some access restrictions apply to the data underlying the findings. Due to ethical and legal restrictions, confidential data are available upon request. To receive anonymized data readers are welcome to contact the corresponding author.

## Author Contribution

T.L..: concept of study design, data acquisition, data post-processing, statistical analysis, drafting of manuscript.

S.K..: data post-processing, statistical analysis, data acquisition, critical revision of manuscript.

A.H. : data post-processing, statistical analysis, critical revision of manuscript.

K.R. : data acquisition, data post-processing, critical revision of manuscript.

C.S.: data acquisition, critical revision of manuscript.

J.S.: data acquisition, critical revision of manuscript.

U.M.: concept of study design, data acquisition, critical revision of manuscript.

M.N.: data post-processing, data acquisition, critical revision of manuscript.

D.H.: data post-processing, data acquisition, critical revision of manuscript.

F.G.: concept of study design, data acquisition, critical revision of manuscript.

M.H.: data post-processing, data acquisition, critical revision of manuscript.

G.S.: concept of study design, data acquisition, critical revision of manuscript.

R.S..: concept of study design, data acquisition, critical revision of manuscript.

S.R.: concept of study design, data acquisition, data post-processing, statistical analysis, drafting of manuscript.

W.H.: concept of study design, data acquisition, data post-processing, statistical analysis, drafting of manuscript.

## Ethics approval

All procedures performed in this study involving human participants were in accordance with the ethical standards with the institutional and the 1964 Helsinki declaration and its later amendments.

## Evidence before this study

We searched PubMed for articles using the search terms "COVID-19“and "aspergillosis“. This search yielded case reports which described invasive pulmonary aspergillosis in critically ill patients with COVID-19. Yet, a systematic prospective evaluation of the risk of invasive pulmonary aspergillosis in a critically ill population with COVID-19 is missing. Also, it remaines to be demonstrated if COVID-19 is independently associated with invasive aspergillosis.

## Added value of this study

This study is to our knowledge the first prospective study performed on the risk for invasive pulmonary aspergillosis in critically ill patients with severe COVID-19 pneumonia. We compared the COVID-19 cohort with matched controls with severe ARDS for the occurrence of invasive pulmonary aspergillosis. With 34% incidence of COVID-19 asscociated invasive pulmonary aspergillosis (CAPA) is higher as in other collectives resulting in an overall ICU mortality rate of 36% as compared to 9.5% in patients without CAPA.

It is also the first study demonstrating that COVID-19 is an independent risk factor for invasive pulmonary aspergillosis by comparison to a COVID-19 negative control cohort.

## Implications of all the available evidence

The independent association of COVID-19 and invasive pulmonary aspergillosis combined with a high mortality rate implicates that standardized screening for COVID-19 associated invasive pulmonary aspergillosis can help to identify patients at risk at an early stage and initiate antimycotic therapy.

